# COVID-19 vaccine hesitancy in the UK: A longitudinal household cross-sectional study

**DOI:** 10.1101/2021.07.09.21260206

**Authors:** Kausik Chaudhuri, Anindita Chakrabarti, Joht Singh Chandan, Siddhartha Bandyopadhyay

**Author notes:** **Corresponding author** Professor Siddhartha Bandyopadhyay. **Contributor ship statement** KC led the quantitative analysis, AC and SB wrote the first draft and JSC provided expertise on the public health implications of the research. All authors reviewed and contributed to the final draft. **Declaration of Interests:** All authors have completed the ICMJE uniform disclosure form at www.icmje.org/coi_disclosure.pdf and declare: no support from any organisation for the submitted work, no financial relationships with any organisations that might have an interest in the submitted work in the previous three years, no other relationships or activities that could appear to have influenced the submitted work. **Ethics approval:** Ethics approval was granted by the University of Essex Ethics Committee for the COVID19 surveys (ETH1920-1271). No additional ethical approval was necessary for this secondary data analysis.

## Abstract

**Background:** The global morbidity and mortality burden of COVID-19 has been substantial, often widening pre-existing inequalities. The approved COVID-19 vaccines have shown great promise in reducing disease transmission and severity of outcomes. However, the success of the COVID-19 vaccine rollout is dependent on public acceptance and willingness to be vaccinated. In this study, we aim to examine how the attitude towards public sector officials and the government impact vaccine willingness with a secondary aim to understand the impact of ethnicity on this relationship.

**Methods:** This cross-sectional study used data from a UK population based longitudinal household survey (Understanding Society COVID-19 study, Understanding Society: the UK Household Longitudinal Study) between April 2020-January 2021. Data from 22421 participants in Waves 6 and 7 of the study were included after excluding missing data. Demographic details in addition to previous survey responses relating to public sector/governmental trust were included in as covariates in the main analyses. A logit model was produced to describe the association between public sector/governmental mistrust and the willingness for vaccination with interaction terms included to account for ethnicity/socio-economic status.

**Findings:** In support of existing literature, we identified those from BAME groups were more likely to be unwilling to take the COVID-19 vaccine. We found that positive opinions towards public sector officials (OR 2.680: 95% CI 1.888 – 3.805) and the UK government (OR 3.400; 95% CI 2.454 - 4.712) led to substantive increase in vaccine willingness. Most notably we identified this effect to be vary across ethnicity and socio-economic status with those from South Asian background (OR 4.513; 95% CI 1.012 - 20.123) the most unwilling to be vaccinated when their trust in public sector officials were affected.

**Interpretation:** These findings suggests that trust in public sector officials may play a key factor in the low vaccination rates particularly seen in at risk groups. Given the additional morbidity/mortality risk posed by COVID-19 to those from lower socio-economic or ethnic minority backgrounds, there needs to be urgent public health action to review how to tailor health promotion advice given to these groups and examine methods to improve trust in public sector officials and the Government.

**Funding:** No funding

**RESEARCH IN CONTEXT:** *Evidence before this study:* A systematic literature search on Pubmed and MedRxiv from database inception to 2^nd^ July 2021 was conducted. The broad terms included were “COVID” OR “SARS-CoV-2*” AND “hesitancy” OR “willingness.” There were no age or language restrictions. We identified numerous observational studies examining prevalence of willingness and hesitancy towards taking the vaccine in a variety of global settings. However, there were fewer studies which examined the reasons behind decisions relating to vaccine hesitancy and in particular in communities relevant to those most at risk in the United Kingdom.

*Added value of this study:* To our knowledge, this is the first attempt to exploring the role of trust in the public sector officials and in the Government with UK COVID-19 vaccination willingness. We found that negative pre-existing opinions around public sector/Government significantly reduced vaccine willingness with an increased effect size noted in those from lower socio-economic and BAME backgrounds.

*Implications of all the available evidence:* Our findings support pre-existing prevalence data suggesting a reduced willingness for vaccination in lower socio-economic/BAME communities. However, our findings build on existing literature by suggesting that trust in public sector officials may play a key factor in the low vaccination rates. Given the additional morbidity/mortality risk posed by COVID-19 to those from lower socio-economic or ethnic minority backgrounds there needs to be urgent public health action to review how to tailor health promotion advice given to these groups and examine methods to improve trust in public sector officials and the Government.

## Background

The COVID-19 pandemic has triggered a concerted global effort to develop and deliver safe efficacious vaccinations at record speed. During the vaccine rollout, the UK has been a leading country as more than three quarters of the population have received at least one dose by early June 2021.^1^ By early April, Public Health England estimated that the vaccines had prevented over 10,000 deaths in people aged over sixty and in Israel, initial evidence demonstrated a marked reduction in the Sars-CoV-2 infection rate and related morbidity and mortality due to vaccination.^2^

Despite the evidenced efficacy, concerns have been raised around existing hesitancy to accept the vaccine which may put the success of the public health initiative at risk.^3^ Within the UK by the 18 February 2021, 34% of 18,855 participants of OCEANS II/III study confirmed that they were either doubtful or strongly unwilling to opt for the vaccine.^4^ Similar mistrust was also noted amongst a higher risk category namely the ‘keyworkers’ with 23.9 % of the 579 keyworkers surveyed confirmed that they are uncertain or will refuse to take the vaccine.^5^ These rates of COVID-19 vaccine hesitancy have remained relatively stable since earlier studies conducted by January 2021 suggesting around 50-60% of individuals would be willing to receive a vaccine.^6,7^ When examining the UK data for markers of inequality, those of 1) black and ethnic minority descendance and 2) lower socioeconomic households or currently unemployed, have indicated even greater rates of hesitancy.^3^ Such findings are like those conducted within the US and France.^8–10^ Concerningly, these groups are at greater risk of transmission of COVID-19 as well as subsequent morbidity and mortality.^11^

Where authors have hypothesised the rationale for the hesitancy in these at-risk groups, uniform messages emanated suggesting that mistrust in the vaccine may correlate with mistrust in public sector governing/health bodies.^3,4,12^ However, this feature has not been explicitly examined within the existing literature, particularly in terms of formal statistical analysis.

Therefore, in this study we aim to examine how the attitude towards public sector officials and the government impact vaccine willingness with a secondary aim to understand the impact of ethnicity and its interaction with the trust variables on this relationship.

## Methods

### Study design and data

This study used data from a UK population based longitudinal household survey (Understanding Society COVID-19 study, Understanding Society: the UK Household Longitudinal Study) between April 2020-January 2021.^13^

The samples are probability samples of postal addresses with slight variations in how the sampling was done across England, Wales and Scotland vs Northern Ireland. In England, Wales and Scotland they are clustered and stratified whereas in Northern Ireland they are unclustered systematic random samples. Northern Ireland and areas in Great Britain with high immigrant and ethnic minority populations were oversampled. The survey consists of all eligible consented individuals aged 16 years and over in eligible households. The survey was undertaken monthly between April-July 2020 and then two monthlies thereafter, with only those who had completed prior surveys eligible for continued entry in the latter months.^14^ The full details for this survey have been described in depth previously.^14^

### Inclusion and outcomes

Although the survey collected data on numerous markers of clinical and social status, in this study we only included those who had been administered/completed the flu and coronavirus vaccine module in November 2020 (wave 6) and in January 2021 (wave 7). The wave 6 survey asked respondents “Imagine that a vaccine against COVID-19 was available for anyone who wanted it. How likely or unlikely would you be to take the vaccine?” The respondent answers options are “very likely, likely, unlikely and very unlikely”. Wave 7 asks the respondents the same question however only to those who had not yet received a COVID vaccine or an appointment for one. In wave 6, a total of 12,035 respondents were eligible but 80 were excluded from our analysis as they either selected the missing/inapplicable/refusal/don’t know responses. For wave 7, out of 11,968 respondents, 23 respondents were excluded due to missing/inapplicable/refusal and 2,109 respondents have received 1^st^ dose, 148 both and 604 respondents received an invitation for vaccination. Combining the two waves (wave 6 and wave 7), we were left with 23,572 respondents.

### Predictor Variables

Although, we only included respondents active and eligible during waves 6 and 7, we utilised covariate data also from the previous waves to track individual respondents over time. Data were taken from previous waves of the COVID-19 survey as well as iterations of the UK Household survey. Doing so provided the following demographic data for use in this study, respondent: 1) age, 2) gender and marital status, 3) ethnicity as per UK census definitions,^15^ 4) attained educational qualifications, 5) employment status, 6) household living arrangements, 7) clinical vulnerability, 8) subjective financial condition, 9) household monthly current income for sensitivity analysis and 10) geographical region. In addition we controlled for the time when survey was conducted. Additionally, in the wave 9 main survey conducted in 2018/19,^14^respondents were asked the following questions: “Public officials don’t care” and “Don’t have a say in what government does”. For both, the respondents’ answers could be “strongly agree/agree, neither agree/disagree, strongly disagree/disagree.” These questions were included in our final dataset as markers of public sector official and government trust.

We use disaggregated ethnicity: South Asian (Indian, Bangladeshi, Pakistani), Black (Black African, Black Caribbean, Other Black), any other Asian, Mixed (white and black Caribbean, white and black African, white and Asian), and other (Arab/any other) with White as the reference category. We include gender (male taking the value of 1), age, educational qualification (degree, A-level/post-secondary, GCSE (General Certificate of Secondary Education), basic (lower than GCSE) with no formal qualification as the base category), employment status (employed, self-employed, both employed & self-employed and none/retired), living arrangement (living with household members aged 70 or older, excluding respondent), clinically vulnerable as identified by NHS (no risk, moderate/high risk), and current subjective financial condition (comfortable/alright, same, and bad). For variables representing markers of public sector official and government trust, the respondents’ answers are “strongly agree/agree, neither agree/disagree, strongly disagree/disagree” (excluding missing/refusal). We use these two variables to create three binary indicators: positive (strongly disagree/disagree), neutral (neither agree/disagree) and negative (strongly agree/agree) and use two of them (positive and neutral) with negative as the base category.

### Statistical analysis

The primary outcome data taken from responses in wave 6 and 7 of the COVID-19 study were re-classified to a vaccine willingness category taking the value of one if the respondents gave a positive response (very likely, likely) and 0 if unlikely and very unlikely. For our combined model, we pooled the data on vaccine willingness category from both rounds. Covariate data were also transformed into binary or categorical groups dependent on the nature of the data.

We use a logit model where the dependent variable (vaccine willingness:) taking the value of 1 and 0 otherwise as follows:

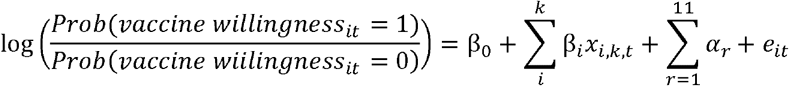

Where *x*_*i,k,t*_, represents covariates/predictor variables for individual i in wave t and is the binary indicator for respondents’ region of residence (there are 12 regions and we use North-East as the reference category). Regression coefficients (*β*_*i*_) were exponentiated and are presented as odds ratios (OR) with corresponding exponentiated 95% confidence intervals (CI) and the p-values (p) for significance with p<0·10 considered to indicate statistical significance. We use robust standard errors to account for some kinds of misspecification allowing for intraclass correlation at the individual level. For the main analyses we analysed two models: Model 1 includes attitude of respondents towards public official and Model 2 includes whether respondents felt they had a say in government. We also ran logit models with interaction terms where we interact attitude of respondents towards public official/ say in government with ethnicity variable (Model 3/Model 4). We use likelihood ratio tests for the interaction model to examine whether their opinion about public officials/government significantly affects the decision of minority communities to get vaccinated. Models were estimated using Stata 15.

### Missing data

Recorded ethnicity was missing for 540 respondents (255 for wave 6 and 285 for wave 7) and these individuals were excluded from our analysis. Ten respondents from each wave were also excluded due to no recorded sex. Educational qualifications were missing for 86 respondents in wave 6 and 82 for wave 7 of COVID -19 survey, who were also excluded. For 14 respondents, geographical location of their residence was unavailable. Further, respondents with missing/inapplicable observations for variables representing as markers of public sector official and government trust were also excluded leaving us with 22421 observations.

## Results

### Study characteristics

During the study period, 22421 respondents were eligible to be included in the final study. Of these the average age was 55 (SD 15.5) with the majority being female (13112/22421 (59.5%). Most respondents were White (20439/22421 (91%)) and in terms of educational status 15878/22421 (71%) were educated to an A-level standard or higher. Although, 10500/22421 (47%) of respondents were currently employed, 16792/22421 (75%) felt financially comfortable/alright. Variables regarding opinion about public officials and caring by government reveal that 4423/22421 (19.7%) and 6012/22424 (26.8%) show positive attitude whereas 8851/22421 (39.5%) and 8084/22424 (36%) display negative attitude. The study characteristics of the sample are presented in Table 1.

**Table 1:** Summary Statistics.

| Variable | Proportion/Mean | SD |
| --- | --- | --- |
| Age | 55.465 | 15.448 |
| Gender |  |  |
| Male | 9309 (0.415) | 0.493 |
| Female | 13112 (0.585) | 0.493 |
| Marital Status |  |  |
| Couple | 15966 (0.712) | 0.453 |
| Ethnicity |  |  |
| South Asian | 931 (0.042) | 0.200 |
| Any other Asian | 226 (0.010) | 0.100 |
| Black | 398 (0.018) | 0.132 |
| Mixed | 320 (0.014) | 0.119 |
| White & Other (ref.) | 20546 (0.916) | 0.277 |
| Educational Qualifications |  |  |
| No qualification dummy (ref.) | 963 (0.043) | 0.203 |
| Basic qualification dummy | 1585 (0.071) | 0.256 |
| GCSE qualification dummy | 3995 (0.178) | 0.383 |
| A-level qualification dummy | 7762 (0.346) | 0.476 |
| Degree dummy | 8116 (0.362) | 0.481 |
| Employment Status |  |  |
| Employed dummy (ref.) | 10500 (0.468) | 0.499 |
| Self-employment dummy | 1685 (0.075) | 0.264 |
| Both Employed/self-employed dummy | 447 (0.020) | 0.140 |
| Retired/others dummy | 9789 (0.437) | 0.496 |
| Household Living Arrangement |  |  |
| No member aged equal/above 70 (ref) | 18692 (0.834) | 0.372 |
| At least 1 member aged equal/above 70 | 3729 (0.166) | 0.372 |
| Clinically Vulnerable Dummy | 9914 (0.442) | 0.497 |
| Subjective Financial Condition |  |  |
| Finance current same as before (ref) | 4665 (0.150) | 0.357 |
| Finance current alright/comfortable | 16792 (0.749) | 0.434 |
| Finance current bad/very bad | 964 (0.043) | 0.203 |
| Opinion about Public Official |  |  |
| Negative opinion (ref) | 8851 (0.395) | 0.489 |
| Neutral opinion | 9147 (0.408) | 0.491 |
| Positive opinion | 4423 (0.197) | 0.398 |
| Opinion about Public Official |  |  |
| Negative opinion (ref) | 8082 (0.360) | 0.480 |
| Neutral opinion | 8328 (0.371) | 0.483 |
| Positive opinion | 6014 (0.268) | 0.443 |

### Main findings

During the study period we found that 19915/22241 (88.8%) of the respondents were willing to take the vaccine. We found that vaccine willingness is positively and significantly associated with age (OR = 1.079 (CI =1.069 - 1.089) in model 1 and 1.077 (1.067 - 1.087) in model 2). People with lower levels of education more likely to be unwilling to take vaccine whereas the clinically vulnerable respondents are more willing [OR: 1.359, CI: 1.046 - 1.765, p-value = 0.02 for Model 1; OR: 1.361, CI: 1.047 - 1.769, p-value = 0.02 for Model 2]. Males and couple are more inclined to get vaccinated compared to females and single. Self-employed are 47 percent against vaccination [OR: 0.471, CI: 0.310 - 0.716, p-value < 0.00 for Model 1; OR: 0.472, CI: 0.310 - 0.718, p-value < 0.00 for Model 2] compared to employed people (base category). Respondents living with 70-plus aged member are more willing to get vaccinated. People from ethnic minority groups shows a varied pattern: Black [OR: 0.004, CI: 0.002 - 0.010, p-value < 0.00 for Model 1 & 2] are least willing, followed by South Asian [OR: 0.106, CI: 0.064 - 0.176, p-value < 0.00 for Model 1 & OR: 0.110, CI: 0.066 - 0.183, p-value < 0.00 for Model 2 respectively]. Note, Black, Asian and Minority Ethnic (BAME) populations are at an increased risk of developing COVID-19 and consequentially more severe outcomes compared to White populations. The odds of vaccine willingness for respondents with better subjective financial well-being is 2.758 (Model 1) and 2.692 (Model 2) times that of respondents with same financial condition; whereas the willingness is significantly lower for financially worse off respondents (OR = 0.493 (CI =0.289 - 0.840) for Model 1 and OR = 0.504 (CI =0.296 - 0.861) for Model 2). Positive/Neutral opinion about care given by public officials increase the odds of vaccine willingness [OR: 1.769, p-value <0.00 (for neutral), and OR: 2.680, p-value <0.00 (for positive), in case of Model 1]. The same correlation with vaccine willingness is corroborated when examining the attitude of respondents towards say in government instead of looking at attitude towards public officials (Model 2).

When examining whether vaccine willingness by ethnic minority differs for those respondents who do not have a negative opinion about care given by public officials/government we observe that all the variables retain their sign and significance as in our original model (Table 2) and respondents without negative opinion about care given by public officials belonging to ethnic minority are more willing to take vaccines. The impact is most significant for two groups, for example, South Asian showing positive attitude [OR: 4.513, CI: 1.012 - 20.123, p-value =0.05] are 4.513 times more willing to get vaccinated compared to the White/Other ethnicity group demonstrating neutral/negative attitude. However, the interaction term between trust and ethnicity is positive for each of the four minority groups.

**Table 2:** Logit regression predictors of Vaccine Willingness.

|  | Model 1<br>OR (p-val) [CI] | Model 2<br>OR (p-val) [CI] |
| --- | --- | --- |
| Age | 1.079***<br>(0.00)<br>[1.069 - 1.089] | 1.077***<br>(0.00)<br>[1.067 - 1.087] |
| Finance current alright/comfortable | 2.758***<br>(0.00)<br>[2.094 - 3.634] | 2.692***<br>(0.00)<br>[2.043 - 3.546] |
| Finance current bad/very bad | 0.493***<br>(0.01)<br>[0.289 - 0.840] | 0.504**<br>(0.01)<br>[0.296 - 0.861] |
| Resp Couple or not | 1.648***<br>(0.00)<br>[1.278 - 2.124] | 1.635***<br>(0.00)<br>[1.269 - 2.106] |
| Male Respondent | 2.411***<br>(0.00)<br>[1.876 - 3.099] | 2.401***<br>(0.00)<br>[1.868 - 3.086] |
| South-Asian | 0.106***<br>(0.00)<br>[0.064 - 0.176] | 0.110***<br>(0.00)<br>[0.066 - 0.183] |
| Any other Asian | 0.266***<br>(0.01)<br>[0.101 - 0.700] | 0.291**<br>(0.01)<br>[0.112 - 0.757] |
| Black | 0.004***<br>(0.00)<br>[0.002 - 0.010] | 0.004***<br>(0.00)<br>[0.002 - 0.010] |
| Mixed | 0.257***<br>(0.00)<br>[0.107 - 0.620] | 0.247***<br>(0.00)<br>[0.102 - 0.596] |
| Clinically Vulnerable | 1.359**<br>(0.02)<br>[1.046 - 1.765] | 1.361**<br>(0.02)<br>[1.047 - 1.769] |
| Basic qualification | 1.372<br>(0.38)<br>[0.673 - 2.797] | 1.376<br>(0.38)<br>[0.675 - 2.803] |
| GCSE qualification | 2.403***<br>(0.01)<br>[1.276 - 4.526] | 2.352***<br>(0.01)<br>[1.251 - 4.423] |
| Alevel/post-secondary | 3.235***<br>(0.00)<br>[1.749 - 5.984] | 3.056***<br>(0.00)<br>[1.654 - 5.646] |
| Degree | 11.154***<br>(0.00)<br>[5.908 - 21.056] | 10.180***<br>(0.00)<br>[5.400 - 19.190] |
| Self-employment | 0.471***<br>(0.00)<br>[0.310 - 0.716] | 0.472***<br>(0.00)<br>[0.310 - 0.718] |
| Both Employed/self-employed | 0.655<br>(0.27) | 0.671<br>(0.30) |
|  | [0.309 - 1.390] | [0.315 - 1.430] |
| Retired/others | 1.093 | 1.091 |
|  | (0.55) | (0.56) |
|  | [0.818 - 1.462] | [0.816 - 1.458] |
| HH with atleast 1 member equal/above 70 | 3.680*** | 3.616*** |
|  | (0.00) | (0.00) |
|  | [2.220 - 6.098] | [2.198 - 5.949] |
| Neutral opinion abt. Public official | 1.769*** |  |
|  | (0.00) |  |
|  | [1.364 - 2.294] |  |
| Positive opinion abt. Public official | 2.680*** |  |
|  | (0.00) |  |
|  | [1.888 - 3.805] |  |
| Neutral opinion abt. say in Govt. |  | 1.654*** |
|  |  | (0.00) |
|  |  | [1.267 - 2.160] |
| Positive opinion abt. say in Govt. |  | 3.400*** |
|  |  | (0.00) |
|  |  | [2.454 - 4.712] |
| *N* | 22,421 | 22,424 |

Based on robust standard error adjusting for clustering at the individual level. \* $p < 0.1$ ; \*\* $p < 0.05$ ; \*\*\* $p < 0.01$

### Sensitivity Analysis

Results reported in Appendix 1 augments Table 3 model by including the monthly income variable. Instead of using income as a continuous variable, we use quartile dummies for income variable (quartile 1 is the lowest and quartile 4 the highest) after adjusting for household size. The odds ratio of all variables along with their significance remains unaltered as reported in Table 3. Higher levels of household monthly income were associated with more willingness to take vaccines.

**Table 3:**
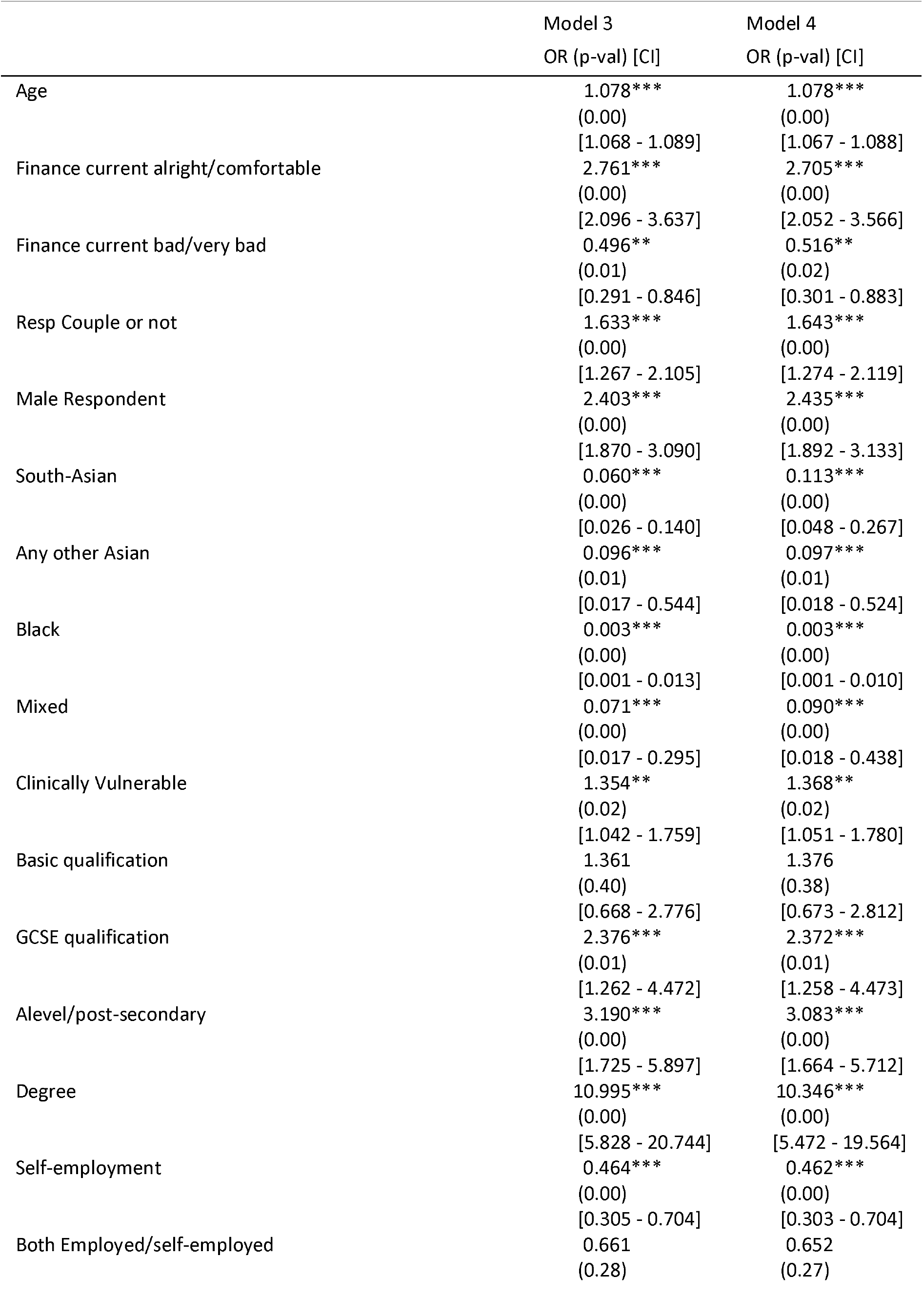
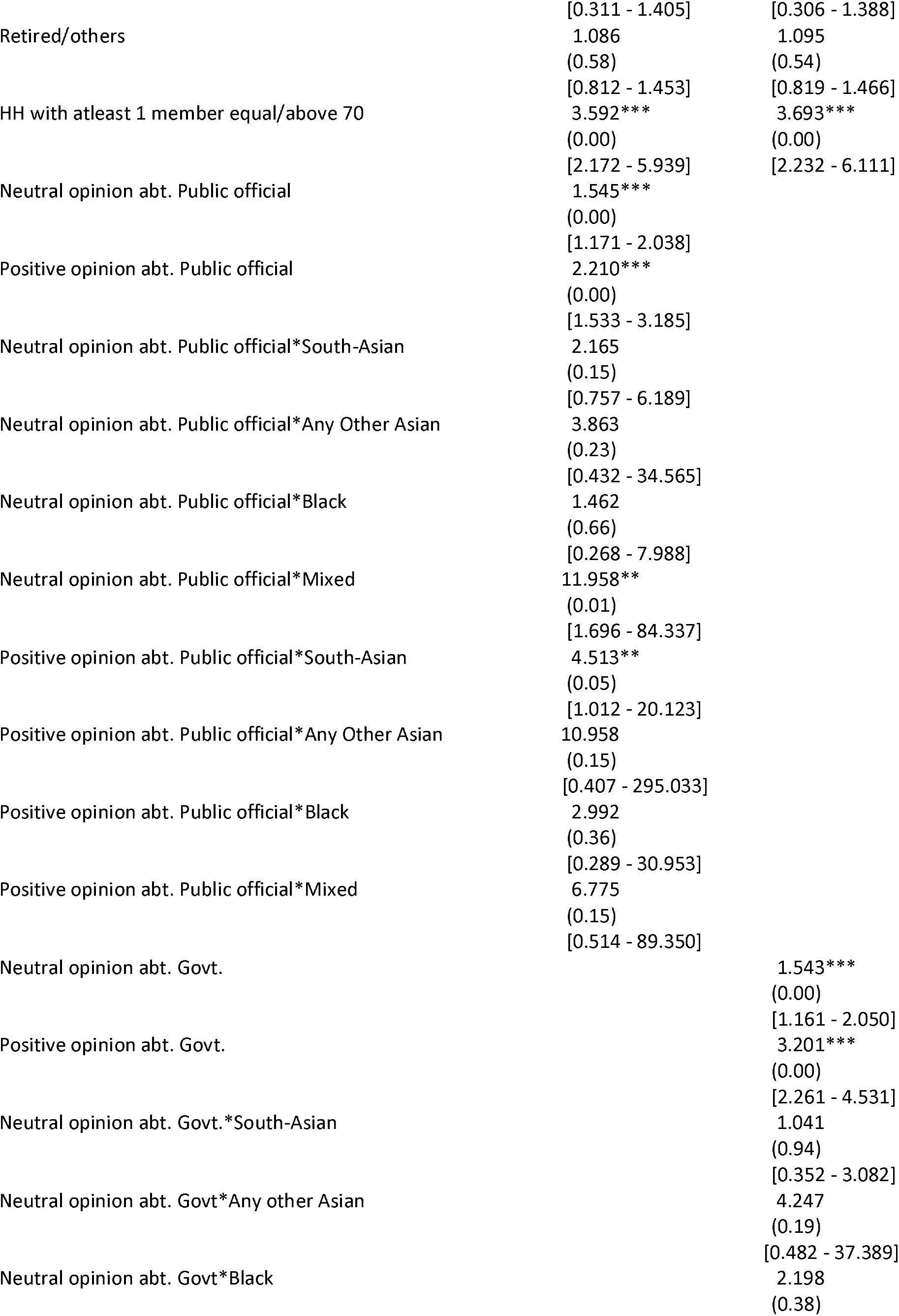
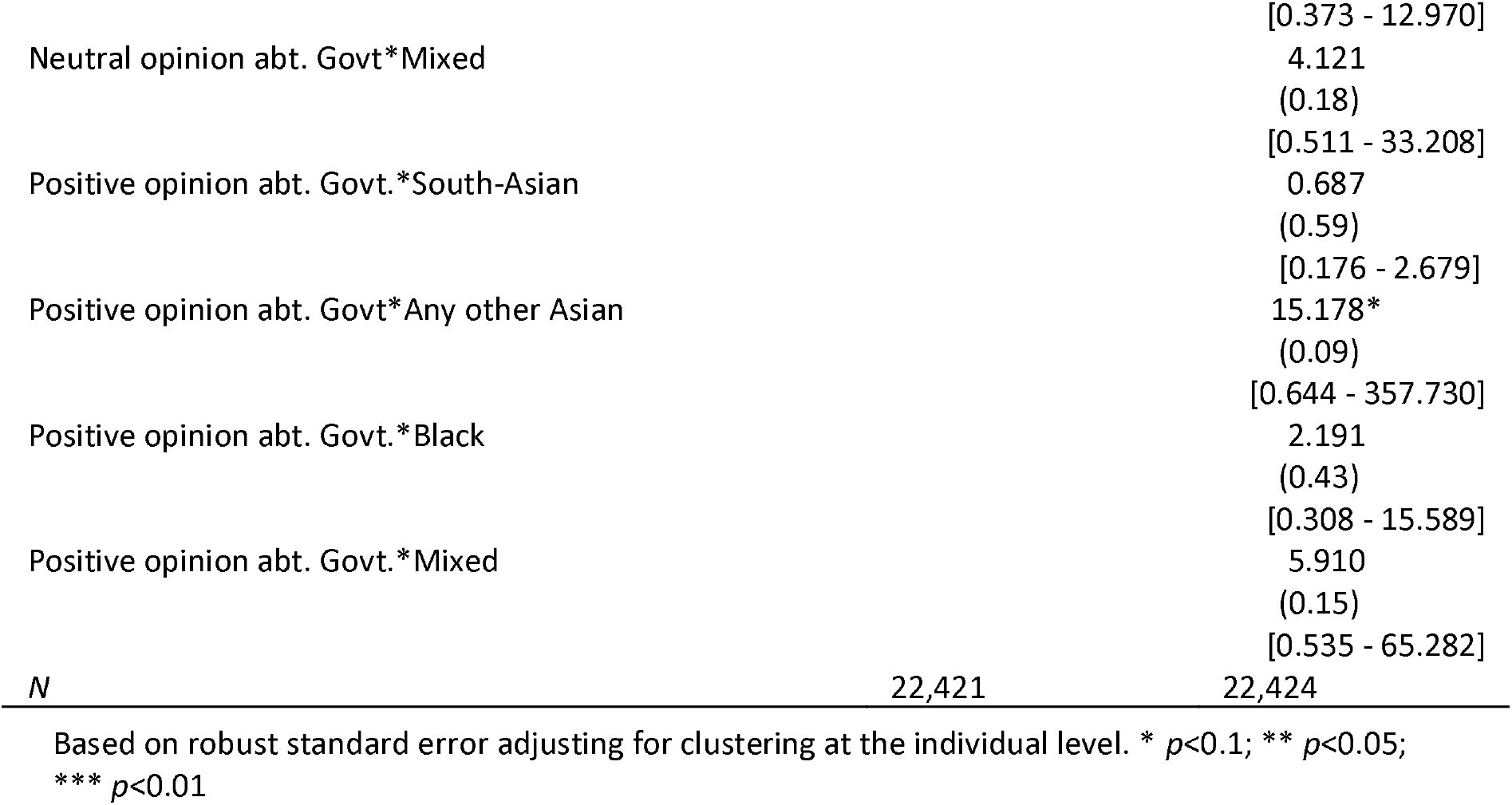
Logit regression predictors of Vaccine Willingness, Interaction Model.

Appendix 2 presents the results disaggregated by age categories (16-34, 34 -49, 50 -64 and 65 plus). The findings show that the odds against vaccine willingness decreases with age. Respondents without negative opinion about care given by public officials belonging to younger age does not depict more willingness to take vaccines. However, people belonging to ethnic minority (combined category of South-Asian/Any Other Asian, Black and Mixed) in the younger age groups not having negative attitude about care given by public officials are more willing to take vaccine with the impact becoming significant for age-group 34-49, for example, ethnic minority in this age-group showing positive attitude [OR: 10.895, CI: 2.239 - 53.009, p-value < 0.00] are almost 11 times more willing to get vaccinated. The same finding is obtained qualitatively with the variable representing respondents’ opinion about government.

## Discussion

In summary we found that COVID-19 vaccine willingness varied substantially based on individual demographics and personal opinions about public sector officials/government. We found that vaccine hesitancy is associated with younger age, being female, not living as couple, lower educational level and income, bad financial subjective wellbeing, belonging to BAME community, and in those who are self-employed. In contrast clinically vulnerable individual and household with adult aged 70 or more portray higher vaccine willingness. It was clear that apart from these demographic factors, the relationship between vaccine willingness was associated with either positive or negative opinions about public officials/government were also associated with higher willingness.

Given the fact that those ethnic minority populations face a higher risk of mortality from COVID, as do those who are from areas with higher levels of deprivation, we might have expected this increased risk to correlate with a higher demand for vaccination in these groups which was not seen in our findings.^11,16^ We identify a crucial factor, namely trust in government which might explain why the BAME population have lower vaccination, something that has been noted as a factor based on a qualitative analysis.^17^ Many in these groups possibly because of discrimination they may have faced tend to mistrust the government which may be exacerbated because of peer effects. This may be an explanatory factor as to why the low uptake we find is highly associated with lower trust. Indeed, among people where trust is not an issue, we find that many ethnic minority populations have a higher willingness to uptake s. Additionally, the relative unwillingness of those who perceive their financial situation to be poor may reflect a higher opportunity cost of time needed to be vaccinated. Conversely, those within the minority ethnic groups who do not hold negative views on the trust questions have a vaccine willingness as shown by the results with the interaction terms. An important point to note is this (lack of) trust is not driven by experiences or perceptions of the government’s Covid management but represents their beliefs on institutional trust as seen by their answers to these questions pre COVID.

As the UK tries to vaccinate its way out of the pandemic, vaccine hesitancy may prove to be a limiting factor that may prevent the full easing of restrictions. Indeed, trust in government has been an important factor that has affected several decisions made by the government on lockdowns and may have potentially affected its timing and intensity.^18^ It has been suggested that lack of trust may well be rooted in historical practices in which minority groups were unethically exploited in medical experiments.^19^ This may also explain why younger sections of the BAME population are less hesitant even though their objective risk of facing death or serious illness from the disease is lower. A public health approach that focuses on increasing uptake of vaccination in these at-risk populations must consider both trust that affects willingness to be vaccinated as well as the differential opportunity cost in terms of the expected time away from work for vaccination (including anticipated time off because of side effects) which makes it costly for certain population segments. Improving institutional trust must be combined with making access easier to improve the coverage of all sections of the population. While our analysis is using data from UK, there is suggestive evidence that some of these factors may be global.

One of the limitations of our study is the use of survey data conducted until January 2021 as people’s vaccination willingness might have changed with arrival of information regarding side-effects of vaccine especially in case AstraZeneca vaccine. Our data reveals that three responses: “I am worried about side effects; I am worried about unknown future effects and I don’t trust vaccines” were the main reasons behind vaccine unwillingness (around 60%). Individuals when being asked for vaccination willingness, the survey did not ask about country of the vaccine manufacturer, type of vaccine individuals are likely to be administered, duration of vaccine immunity and place of vaccine administration. During the two rounds of survey, the UK was going through the second wave of Covid-19 and willingness to take vaccine might have changed in response to increasing numbers of infections/deaths. Also, the respondents may not have been aware of vaccine efficacy outside clinical trials especially in context of hospital admissions/severe illness and this could impact COVID-19 vaccine willingness.

In order to begin the recovery phase of the COVID-19 pandemic, there is an urgency to implement strong and successful global vaccine programmes. However, vaccine hesitancy may derail any intention to do so. Our findings have confirmed previous findings suggesting those from lower socio-economic and minority ethnic communities have the highest rates of vaccine hesitancy. Upon further examination it is clear that this relationship is mediated by trust in public sector officials or the Government. Therefore, urgent action is needed to promote public health messaging to build trust to encourage improved uptake particularly in groups who are most at risk of negative clinical consequences of COVID-19.

## Data Availability

The data is publicly available from the link provided

https://www.understandingsociety.ac.uk/documentation

## Figures and tables

**Figure 1:**
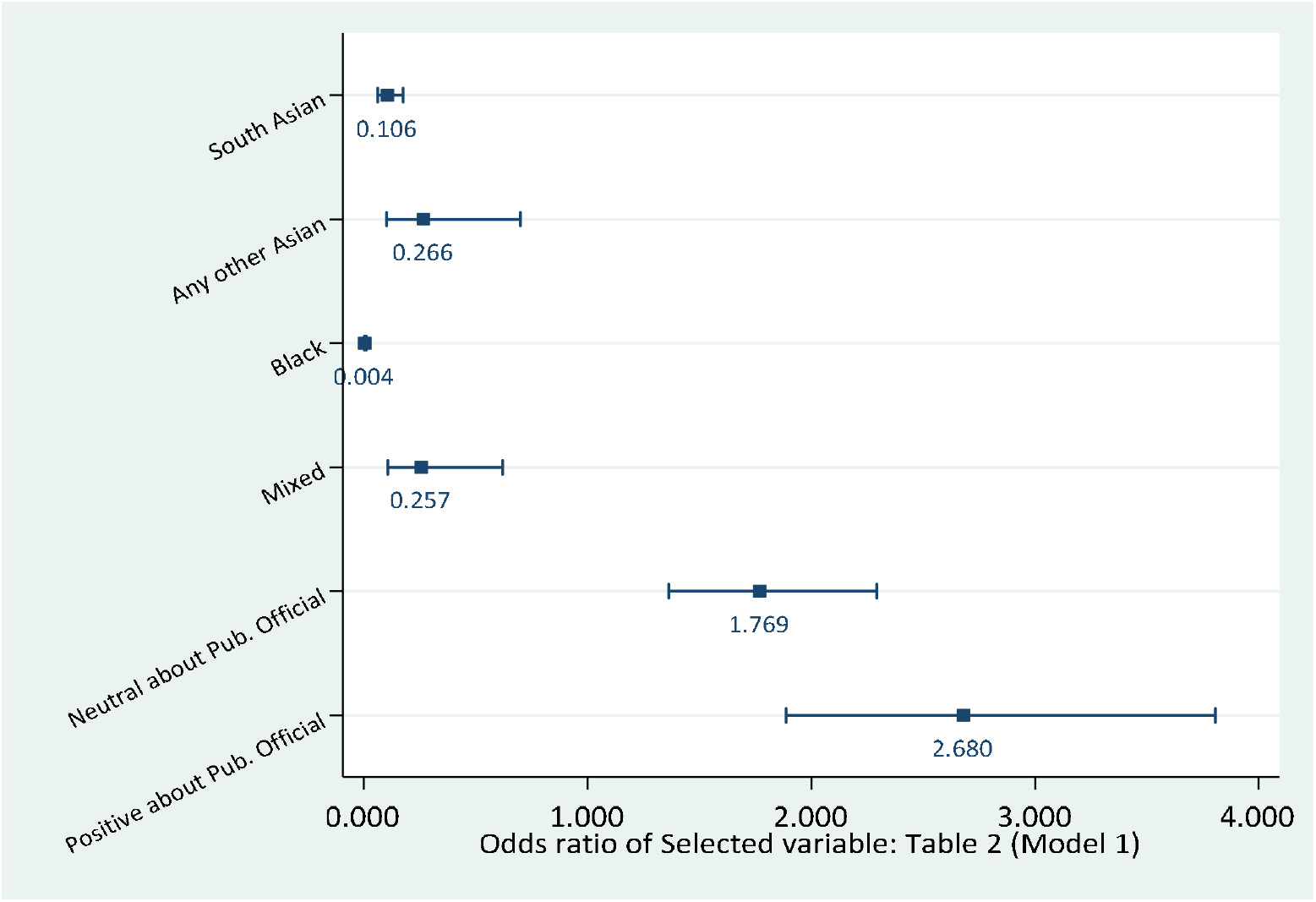
Ethnicity and Opinion about Care given by Public Official.

**Figure 2:**
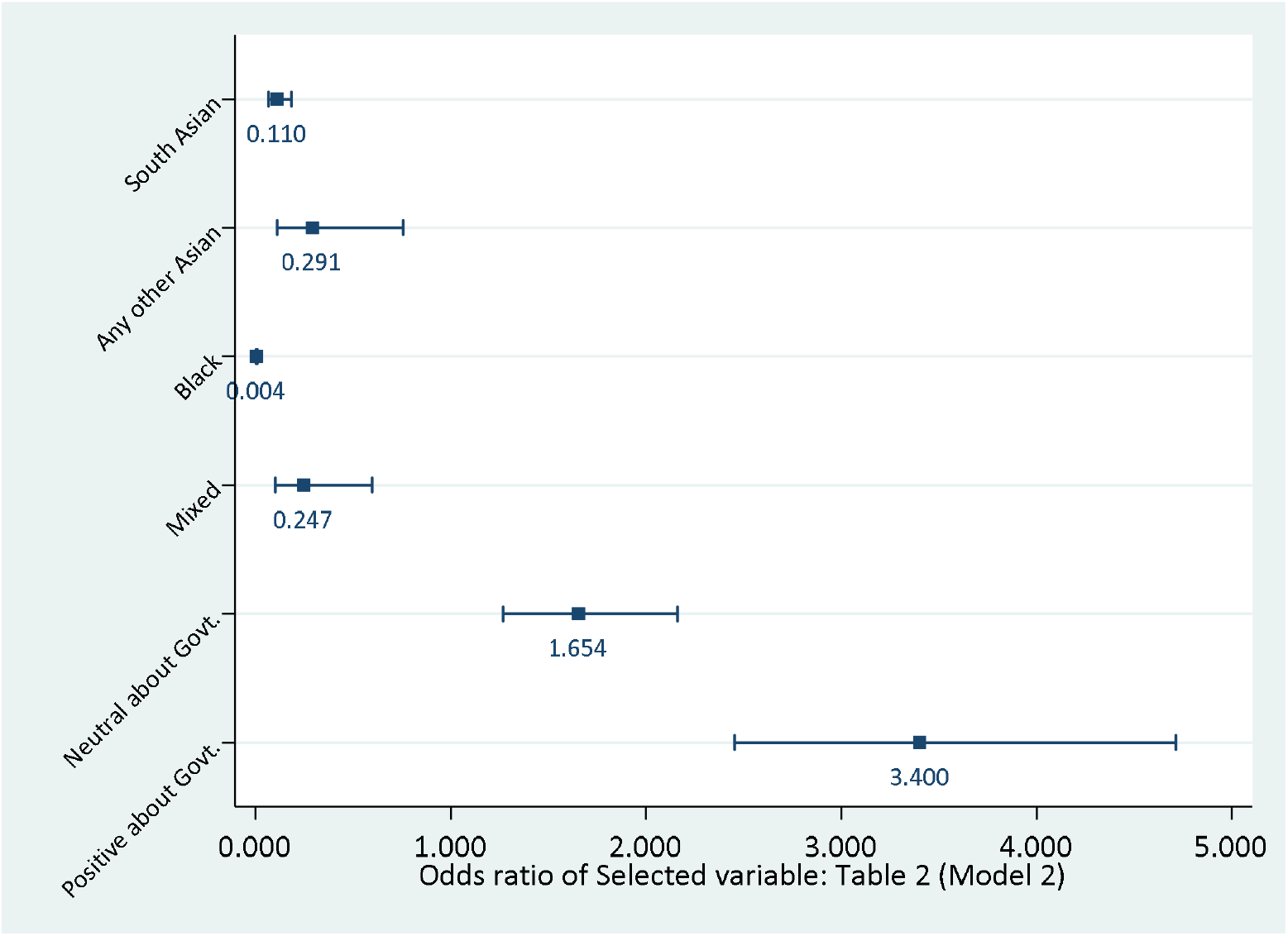
Ethnicity and Opinion about Say in Government Activities.

