## Appendix for "COVID-19 vaccine hesitancy in the UK: A longitudinal household cross-sectional study"

**Appendix 1: Logit regression predictors of Vaccine Willingness, Interaction Model with Income**

|  | Model 3 with income added  OR (p-val) [CI] | Model 4 with income added  OR (p-val) [CI] |
| --- | --- | --- |
| Age | 1.075*** | 1.075*** |
|  | (0.00) | (0.00) |
|  | [1.063 - 1.088] | [1.062 - 1.088] |
| Finance current alright/comfortable | 2.514*** | 2.452*** |
|  | (0.00) | (0.00) |
|  | [1.796 - 3.518] | [1.751 - 3.433] |
| Finance current bad/very bad | 0.585* | 0.610 |
|  | (0.09) | (0.12) |
|  | [0.313 - 1.093] | [0.325 - 1.146] |
| Resp Couple or not | 1.988*** | 1.981*** |
|  | (0.00) | (0.00) |
|  | [1.470 - 2.689] | [1.465 - 2.678] |
| Male Respondent | 2.445*** | 2.461*** |
|  | (0.00) | (0.00) |
|  | [1.812 - 3.299] | [1.823 - 3.322] |
| South Asian | 0.074*** | 0.137*** |
|  | (0.00) | (0.00) |
|  | [0.027 - 0.201] | [0.049 - 0.379] |
| Any other Asian | 0.105** | 0.066*** |
|  | (0.01) | (0.01) |
|  | [0.017 - 0.640] | [0.010 - 0.446] |
| Black | 0.004*** | 0.003*** |
|  | (0.00) | (0.00) |
|  | [0.001 - 0.022] | [0.001 - 0.015] |
| Mixed | 0.063*** | 0.163** |
|  | (0.00) | (0.05) |
|  | [0.011 - 0.351] | [0.027 - 0.993] |
| Clinically Vulnerable Dummy =1 | 1.323* | 1.318* |
|  | (0.09) | (0.09) |
|  | [0.961 - 1.821] | [0.957 - 1.815] |
| Basic qualification | 0.595 | 0.589 |
|  | (0.24) | (0.23) |
|  | [0.252 - 1.407] | [0.248 - 1.396] |
| GCSE qualification | 1.240 | 1.208 |
|  | (0.59) | (0.63) |
|  | [0.572 - 2.688] | [0.557 - 2.622] |
| Alevel/post-secondary qualification | 1.716 | 1.638 |
|  | (0.16) | (0.20) |
|  | [0.809 - 3.639] | [0.771 - 3.480] |
| Degree | 5.955*** | 5.452*** |
|  | (0.00) | (0.00) |
|  | [2.747 - 12.909] | [2.510 - 11.846] |
| Self-employment | 0.465*** | 0.470*** |
|  | (0.00) | (0.00) |
|  | [0.277 - 0.781] | [0.279 - 0.793] |
| Both Employed/self-employed | 0.631 | 0.618 |
|  | (0.31) | (0.28) |
|  | [0.262 - 1.522] | [0.256 - 1.490] |
| Retired/others | 1.261 | 1.254 |
|  | (0.20) | (0.21) |
|  | [0.884 - 1.800] | [0.879 - 1.790] |
| HH with at least 1 member equal/above 70 | 3.734*** | 3.724*** |
|  | (0.00) | (0.00) |
|  | [2.040 - 6.835] | [2.040 - 6.798] |
| Neutral opinion abt. Public official | 1.332* |  |
|  | (0.09) |  |
|  | [0.958 - 1.854] |  |
| Positive opinion abt. Public official | 1.974*** |  |
|  | (0.00) |  |
|  | [1.290 - 3.022] |  |
| Neutral opinion abt. Public official*South-Asian | 4.050** |  |
|  | (0.03) |  |
|  | [1.116 - 14.695] |  |
| Neutral opinion abt. Public official*Any other Asian | 2.370 |  |
|  | (0.46) |  |
|  | [0.240 - 23.419] |  |
| Neutral opinion abt. Public official*Black | 1.965 |  |
|  | (0.52) |  |
|  | [0.253 - 15.246] |  |
| Neutral opinion abt. Public official*Mixed | 12.888** |  |
|  | (0.03) |  |
|  | [1.305 - 127.283] |  |
| Positive opinion abt. Public official*South-Asian | 7.672** |  |
|  | (0.04) |  |
|  | [1.148 - 51.275] |  |
| Positive opinion abt. Public official*Any other Asian | 10.284 |  |
|  | (0.25) |  |
|  | [0.197 - 36.336] |  |
| Positive opinion abt. Public official*Black | 2.240 |  |
|  | (0.60) |  |
|  | [0.109 - 45.863] |  |
| Positive opinion abt. Public official*Mixed | 4.827 |  |
|  | (0.30) |  |
|  | [0.248 - 94.054] |  |
| Neutral opinion abt. Govt. |  | 1.356* |
|  |  | (0.08) |
|  |  | [0.967 - 1.902] |
| Positive opinion abt. Govt. |  | 2.770*** |
|  |  | (0.00) |
|  |  | [1.846 - 4.157] |
| Neutral opinion abt. Govt*South-Asian |  | 1.912 |
|  |  | (0.34) |
|  |  | [0.507 - 7.211] |
| Neutral opinion abt. Govt*Any other Asian |  | 4.947 |
|  |  | (0.19) |
|  |  | [0.453 - 54.055] |
| Neutral opinion abt. Govt*Black |  | 2.138 |
|  |  | (0.48) |
|  |  | [0.259 - 17.671] |
| Neutral opinion abt. Govt*Mixed |  | 2.111 |
|  |  | (0.55) |
|  |  | [0.186 - 23.914] |
| Positive opinion abt. Govt.*South-Asian |  | 1.231 |
|  |  | (0.81) |
|  |  | [0.236 - 6.433] |
| Positive opinion abt. Govt.*Any other Asian |  | 22.877 |
|  |  | (0.11) |
|  |  | [0.471 - 41.037] |
| Positive opinion abt. Govt.*Black |  | 4.271 |
|  |  | (0.24) |
|  |  | [0.386 - 47.244] |
| Positive opinion abt. Govt.*Mixed |  | 0.925 |
|  |  | (0.95) |
|  |  | [0.066 - 12.940] |
| Income quartile dummy 2 | 1.291 | 1.274 |
|  | (0.11) | (0.13) |
|  | [0.942 - 1.768] | [0.929 - 1.747] |
| Income quartile dummy 3 | 1.822*** | 1.777*** |
|  | (0.00) | (0.00) |
|  | [1.289 - 2.575] | [1.258 - 2.512] |
| Income quartile dummy 4 | 2.580*** | 2.512*** |
|  | (0.00) | (0.00) |
|  | [1.740 - 3.825] | [1.696 - 3.722] |
| *N* | 16,412 | 16,414 |

Based on robust standard error adjusting for clustering at the individual level. * *p*<0.1; ** *p*<0.05; *** *p*<0.01.

**Appendix 2: Logit regression predictors of Vaccine Willingness, Interaction Model (disaggregated age)**

|  | Model 1  OR (p-val) [CI] | Model 2  OR (p-val) [CI] |
| --- | --- | --- |
| Age 16-34 | 0.075*** | 0.051*** |
|  | (0.00) | (0.00) |
|  | [0.038 - 0.150] | [0.025 - 0.106] |
| Age 35-49 | 0.089*** | 0.078*** |
|  | (0.00) | (0.00) |
|  | [0.047 - 0.166] | [0.040 - 0.150] |
| Age 50-64 | 0.319*** | 0.239*** |
|  | (0.00) | (0.00) |
|  | [0.179 - 0.570] | [0.130 - 0.440] |
| Finance current alright/comfortable | 2.754*** | 2.739*** |
|  | (0.00) | (0.00) |
|  | [2.086 - 3.635] | [2.074 - 3.619] |
| Finance current bad/very bad | 0.518** | 0.531** |
|  | (0.02) | (0.02) |
|  | [0.301 - 0.892] | [0.308 - 0.915] |
| Resp Couple or not | 1.977*** | 1.953*** |
|  | (0.00) | (0.00) |
|  | [1.527 - 2.561] | [1.508 - 2.530] |
| Male Respondent | 2.561*** | 2.566*** |
|  | (0.00) | (0.00) |
|  | [1.992 - 3.292] | [1.994 - 3.302] |
| Clinically Vulnerable | 1.439*** | 1.443*** |
|  | (0.01) | (0.01) |
|  | [1.101 - 1.879] | [1.104 - 1.887] |
| Basic qualification | 1.439 | 1.447 |
|  | (0.33) | (0.32) |
|  | [0.695 - 2.981] | [0.699 - 2.993] |
| GCSE | 2.298** | 2.280** |
|  | (0.01) | (0.01) |
|  | [1.210 - 4.365] | [1.201 - 4.328] |
| Alevel/post-secondary | 3.073*** | 2.952*** |
|  | (0.00) | (0.00) |
|  | [1.648 - 5.728] | [1.583 - 5.503] |
| Degree | 11.470*** | 10.603*** |
|  | (0.00) | (0.00) |
|  | [6.027 - 21.831] | [5.569 - 20.190] |
| Self-employment | 0.497*** | 0.510*** |
|  | (0.00) | (0.00) |
|  | [0.324 - 0.762] | [0.332 - 0.783] |
| Both Employed/self-employed | 0.642 | 0.666 |
|  | (0.26) | (0.30) |
|  | [0.298 - 1.382] | [0.308 - 1.438] |
| Retired/others | 1.075 | 1.095 |
|  | (0.64) | (0.56) |
|  | [0.793 - 1.458] | [0.806 - 1.486] |
| HH with atleast 1 member equal/above 70 | 3.779*** | 3.857*** |
|  | (0.00) | (0.00) |
|  | [2.229 - 6.406] |  |
| Neutral opinion abt. Public official | 2.598** |  |
|  | (0.01) |  |
|  | [1.247 - 5.413] |  |
| Positive opinion abt. Public official | 2.730** |  |
|  | (0.04) |  |
|  | [1.038 - 7.177] |  |
| Neutral opinion abt. Public official |  | 1.590 |
|  |  | (0.22) |
|  |  | [0.757 - 3.338] |
| Positive opinion abt. Public official |  | 2.362* |
|  |  | (0.05) |
|  |  | [0.989 - 5.641] |
| Neutral opinion abt. Public official*Age 16-34 | 0.358** |  |
|  | (0.04) |  |
|  | [0.134 - 0.955] |  |
| Neutral opinion abt. Public official*Age 34-49 | 0.639 |  |
|  | (0.32) |  |
|  | [0.264 - 1.545] |  |
| Neutral opinion abt. Public official*Age 50-64 | 0.627 |  |
|  | (0.28) |  |
|  | [0.269 - 1.466] |  |
| Positive opinion abt. Public official*Age 16-34 | 0.378 |  |
|  | (0.13) |  |
|  | [0.108 - 1.320] |  |
| Positive opinion abt. Public official*Age 34-49 | 0.672 |  |
|  | (0.50) |  |
|  | [0.214 - 2.114] |  |
| Positive opinion abt. Public official*Age 50-64 | 1.408 |  |
|  | (0.56) |  |
|  | [0.440 - 4.501] |  |
| Minority | 0.031*** | 0.042*** |
|  | (0.00) | (0.00) |
|  | [0.016 - 0.059] | [0.022 - 0.080] |
| Neutral opinion abt. Public official*Age 16-34*Minority | 2.244 |  |
|  | (0.24) |  |
|  | [0.576 - 8.733] |  |
| Neutral opinion abt. Public official*Age 34-49*Minority | 2.587* |  |
|  | (0.08) |  |
|  | [0.887 - 7.550] |  |
| Neutral opinion abt. Public official*Age 50-64*Minority | 1.786 |  |
|  | (0.28) |  |
|  | [0.630 - 5.067] |  |
| Positive opinion abt. Public official*Age 16-34*Minority | 5.616 |  |
|  | (0.16) |  |
|  | [0.496 - 63.622] |  |
| Positive opinion abt. Public official*Age 34-49*Minority | 10.895*** |  |
|  | (0.00) |  |
|  | [2.239 - 53.009] |  |
| Positive opinion abt. Public official*Age 50-64*Minority | 1.365 |  |
|  | (0.73) |  |
|  | [0.233 - 7.986] |  |
| Neutral opinion abt. Govt.*Age 16-34 |  | 0.743 |
|  |  | (0.56) |
|  |  | [0.273 - 2.025] |
| Neutral opinion abt. Govt.*Age 34-49 |  | 0.894 |
|  |  | (0.81) |
|  |  | [0.365 - 2.193] |
| Neutral opinion abt. Govt.*Age 50-64 |  | 1.210 |
|  |  | (0.66) |
|  |  | [0.512 - 2.859] |
| Positive opinion abt. Govt.*Age 16-34 |  | 1.140 |
|  |  | (0.82) |
|  |  | [0.358 - 3.625] |
| Positive opinion abt. Govt.*Age 34-49 |  | 1.112 |
|  |  | (0.84) |
|  |  | [0.390 - 3.168] |
| Positive opinion abt. Govt.*Age 50-64 |  | 1.955 |
|  |  | (0.21) |
|  |  | [0.687 - 5.559] |
| Neutral opinion abt. Govt.*Age 16-34*Minority |  | 1.625 |
|  |  | (0.50) |
|  |  | [0.400 - 6.596] |
| Neutral opinion abt. Govt.*Age 34-49*Minority |  | 1.745 |
|  |  | (0.33) |
|  |  | [0.575 - 5.295] |
| Neutral opinion abt. Govt.*Age 50-64*Minority |  | 1.688 |
|  |  | (0.35) |
|  |  | [0.559 - 5.094] |
| Positive opinion abt. Govt.*Age 16-34*Minority |  | 0.898 |
|  |  | (0.91) |
|  |  | [0.127 - 6.348] |
| Positive opinion abt. Govt.*Age 34-49*Minority |  | 4.647** |
|  |  | (0.03) |
|  |  | [1.177 - 18.356] |
| Positive opinion abt. Govt.*Age 50-64*Minority |  | 0.585 |
|  |  | (0.49) |
|  |  | [0.126 - 2.713] |
| *N* | 22,421 | 22,424 |

Based on robust standard error adjusting for clustering at the individual level. * *p*<0.1; ** *p*<0.05; *** *p*<0.01. Minority includes South-Asian, Any other Asian, Black and Mixed.
